# Real-life evaluation of a rapid antigen test (DPP® SARS-CoV-2 Antigen) for COVID-19 diagnosis of primary healthcare patients, in the context of the Omicron-dominant wave in Brazil

**DOI:** 10.1101/2022.08.02.22278277

**Authors:** Matheus Filgueira Bezerra, Lilian Caroliny Amorim Silva, Rômulo Pessoa-e-Silva, Gisele Lino Soares, Filipe Zimmer Dezordi, Gustavo Barbosa de Lima, Raul Emídio de Lima, Tulio L. Campos, Cassia Docena, Anderson Bruno de Oliveira, Maira Galdino da Rocha Pitta, Francisco de Assis da Silva Santos, Michelly Pereira, Gabriel Luz Wallau, Marcelo Henrique Santos Paiva

## Abstract

Rapid antigen tests play an important role in the monitoring and mitigation of the COVID-19 pandemic, as it provides an easy, fast and efficient diagnosis with minimum infrastructure requirements. However, as new variants of concern continue to emerge, mutations in the virus genome may impair the recognition of the mutated antigen by the tests. Therefore, it is essential to re-assess the test’s sensitivity as the virus mutation profile undergoes significant changes. Here, we prospectively accessed the performance of the DPP® SARS-CoV-2 Antigen test in the context of an omicron-dominant real-life setting. We evaluated 347 unselected individuals (all-comers) from a public testing center in Brazil, performing the rapid antigen test diagnosis at point-of-care with fresh samples. The combinatory result from two distinct RT-qPCR methods was employed as reference and 13 samples with discordant PCR results were excluded. The assessment of the rapid test in 67 PCR-positive and 265 negative samples revealed an overall sensitivity of 80.5%, specificity of 99.2% and positive/negative predictive values higher than 95%. However, we observed that the sensitivity was dependent on the viral load (sensitivity in Ct<31 = 93.7%; Ct>31 = 47.4%). Furthermore, we were able to confirm that the positive samples evaluated in the study were Omicron (BA.1/BA.1.1) by whole-genome sequencing (n=40) and multiplex RT-qPCR (n=17). Altogether, the data obtained from a real-life prospective cohort supports that the rapid antigen test sensitivity for the Omicron remains high and underscores the reliability of the test for COVID-19 diagnosis in a setting with high disease prevalence and limited PCR testing capability.

## INTRODUCTION

Rapid antigen tests are a powerful tool in the mitigation of the COVID-19 pandemic since it does not require complex laboratory infrastructure and drastically reduces the time to results release. Together, these features increase the testing capacity of the healthcare system and reduce the time to identify the infected individuals, allowing a more effective blockage in the chain of transmission (Koo et al., 2022). These tests are particularly useful in scenarios of massive community spread of the virus, with high demands for PCR tests and the need for prompt identification of infected individuals. However, much of the currently available antigen-capture tests are based on the nucleocapsid aminoacid sequences from the early SARS-CoV-2 lineages circulating in 2020 (Raich-Regué et al., 2022). As of March 2022, those early lineages have been fully displaced by multiple waves produced by new COVID-19 Variants of Concern (VOCs) and the new cases in Brazil are caused mostly by the highly mutated Omicron variant and its sublineages (http://www.genomahcov.fiocruz.br/dashboard-en/).

Therefore, it is not clear yet whether multiple mutations accumulated by these variants in the target nucleocapsid antigen affects or not the binding affinity of antibodies employed in the antigen-capture rapid tests and hence COVID-19 diagnosis. Therefore, we prospectively accessed the performance of a rapid antigen test widely used in Brazilian public healthcare assistance, in the context of an omicron-dominant wave real-life setting.

## METHODS

### Samples

The study design consisted of a prospective collection of 347 unselected individuals (all-comers) that reached the public COVID-19 Testing Center in the city of Caruaru (Pernambuco – Brazil). Sample collection was performed between February 15^th^ and March 29^th^, 2022, during the decline of the outbreak caused by the spread of Omicron (Pangolin BA.1 lineage) in the region. The patients setting included symptomatic individuals, contactants of confirmed cases and asymptomatic individuals that needed testing for specific reasons such as traveling or access to public events. All patients enrolled were first asked to be part of the study and only then, were simultaneously collected with two swabs: one for the PCR test and the other for immediate application of a rapid antigen test.

### Ethics Statement

The study was approved by the local Ethics Committee (CEP-CCS/UFPE, CAAE 31093420.4.0000.5208) and all the participants voluntarily accepted to be part of the study and signed the written consent.

### Rapid antigen test

The DPP® SARS-CoV-2 Antigen test is produced by Chembio Diagnostic Systems (Medford, USA) and is registered in Brazil by the Instituto de Tecnologia em Imunobiológicos, Biomanguinhos (Rio de Janeiro, Brazil), with the name TR DPP® COVID-19 AG. It consists of a Dual Path Platform system (DPP®) antigen assay that detects the SARS-CoV-2 nucleocapsid antigen from nasopharyngeal samples.

The rapid tests were performed *in loco*, immediately after sample collection, following the manufacturer’s instructions. In short, collected swabs were mixed with the sampling buffer in collection tubes and transferred to the test platform. After five minutes, the buffer was also added in the second well. After 20 minutes of incubation at room temperature, the test was read by two observers and results were promptly reported back to the patients.

### Molecular diagnosis of COVID-19

Swabs from all 347 patients were processed in the molecular diagnosis laboratory from the Suely-Galdino Therapeutic Innovation Research Center (NUPIT-SG) and results were reported back to the patients. In order to increase the precision of PCR diagnosis, here used as the reference method to evaluate the rapid test, all samples were simultaneously tested with two RT-qPCR diagnosis-validated assays, as described below.

Total RNA extraction was carried out using the Maxwell® RSC Viral Total Nucleic Acid Purification Kit with a Maxwell® RSC (Promega, WI, USA), with a sample input of 200 µL. The first PCR was performed using the GoTaq®C Probe 1-Step-RT-qPCR System (Promega) and the 2019-nCoV_N1/N2/RP primers/probes from CDC (Centers for Disease Control and Prevention, Atlanta, USA). The second PCR was performed using the Duplex SARS-CoV-2 E/RP Molecular Kit (Bio-Manguinhos/FIOCRUZ, Rio de Janeiro, Brazil). Both assays were executed and interpreted following the manufacturer instructions. All assays were run in a QuantStudio™ 5 Real-Time PCR System 96-well, and results were analyzed with the QuantStudio™ 3 & 5 qPCR Data Analysis Software (Applied Biosystems®, Foster City, USA).

The combinatory result from the two PCRs was assigned as “valid” when it had two concordant results (positive/positive or negative/negative), or one valid (positive or negative) result combined with one inconclusive. Samples that had divergent PCR results (positive/negative) or two inconclusive results were assigned as “invalid” and excluded from the further analysis (Figure 1).

**Figure 1.**
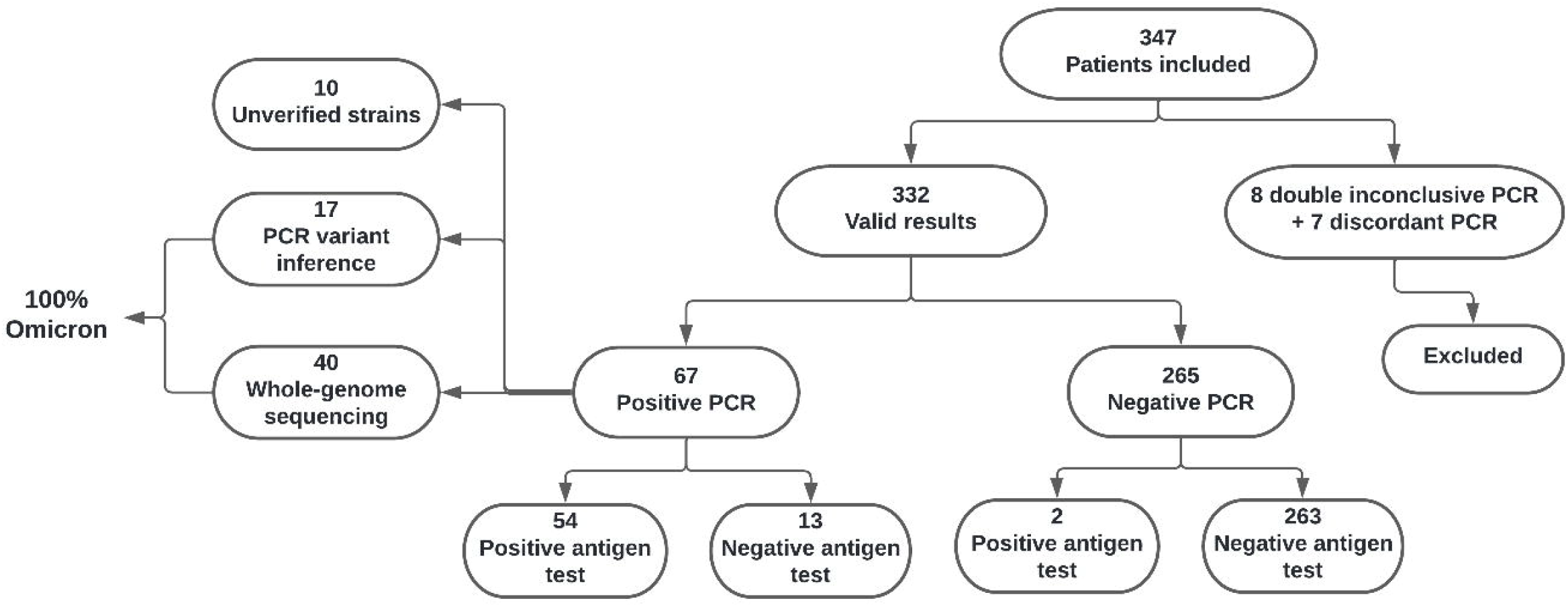
Study design and workflow. The organogram displays the amount of samples evaluated and their distribution according to inclusion criteria, diagnostic tests results and sequencing.

### SARS-CoV-2 genome sequencing

Nasopharyngeal samples with Ct values ≤ 29 in RT-qPCR tests were considered eligible for genome sequencing. To generate cDNA and amplify the SARS-CoV-2 genome library, we employed the ARTIC V4 primers (Loman, 2021), inserting three complementary sets of primers according to Naveca et al. (2022). Whole genome amplicons were then prepared for sequencing using the Illumina COVIDSeq Test (Illumina, San Diego, CA, USA), following the manufacturer’s instructions. MiSeq high throughput sequencer coupled with MiSeq Reagent kit V3 -paired-end 150 cycles flow cell was used for sequencing.

### Genome assembly, annotation and phylogenomic data analysis

Consensus genomes of sequenced samples were assembled using ViralFlow v.0.0.6 (Dezordi, 2022) using 5 reads as minimum coverage depth threshold. Lineages were determined using pangolin v.4.0.6 (O’Toole, 2021) and consensus genomes were then submitted to GISAID (SHU, 2017). We performed a multiple sequence alignment based on the Omicron variant (BA.1 and BA.1* sublineages) using a focal-contextual strategy of Nextstrain (v.3.0.3) (Aksamentov, 2021). This alignment was enriched with genomes from Pernambuco state (focal) and genomes from Brazil and the World that were sorted based on location and collection date and then selected based on the proximity with genomes from Pernambuco (contextual). The focal-contextual strategy was based on selecting up to 3000 genomes from Pernambuco state, 250 from Brazil (excluding Pernambuco), 150 from South America (excluding Brazil) and 50 from the rest of globe (Supplementary File 1). The genomes from Pernambuco are not grouped, the genomes from other groups (Brazil, South America, Globe) are grouped based on location and collection date, and the priority for sequence choice is based on the genetic proximity with Pernambuco sequences. Moreover, 50 sequences are requested as outgroups, from B.1.1 (B.1.1 parent lineage) and B.1.1.529 (BA.1 parent lineage) lineages.

To reconstruct the phylogenetic tree, we first aligned the 40 genomes generated in this study against the reference alignment using MAFFT (Katoh, 2013). Then, we performed a maximum likelihood reconstruction using IQ-TREE2 (Minh, 2020) with both aLRT and ultrafast bootstrap (Hoang, 2018) and selected the substitution model with ModelFinder (Kalyaanamoorthy, 2017). The reconstructed tree was annotated using iTOL (Letunic, 2019) considering the PANGOLIN (Phylogenetic Assignment of Named Global Outbreak Lineages) lineage information.

### VOC inference PCR assay

For cases with lower viral loads, the SARS-CoV-2 lineages were inferred using a multiplex RT-qPCR, and a panel of primers and probes for detection of specific mutations was used. This molecular tool is based on the mutational profile of the VOCs, as following: Alpha (B.1.1.7, B.1.258, B.1.525 – S.delH69V70), Gamma (P.1 – S.K417T.AAG.ACG), Delta (B.1.617.2 – S.P681R.CCT.CGT) and Omicron (B.1.1.529 – S.G339D.GGT_GAT). All probes and the TaqPath™ 1-Step RT-qPCR Master Mix, CG were acquired from Thermo Fisher Scientific (Waltham, MA, USA), and used according to the manufacturer’s instructions. The assays were run in a QuantStudio™ 5 Real-Time PCR System 96-well (Applied Biosystems®, Foster City, USA).

### Epidemiological and clinical data

Data regarding COVID-19 confirmed cases in the city of Caruaru state were obtained from the coronavirus website of the Brazilian Ministry of Health (https://covid.saude.gov.br/). The clinical data were obtained from the patients’ files at the Caruaru Testing Center.

### Statistical Analysis

The combinatory result of the two PCR methods was used as the gold standard to calculate the DPP® COVID-19 AG performance rates. Sensitivity, specificity, accuracy and confidence intervals were calculated using the medcalc platform (https://www.medcalc.org).

Scatterplots and correlation analysis were calculated using the GraphPad Prism version 8 software. Spearman test was used to measure the correlation between Cts from the two PCR methods and Mann-Whitney test was used to compare Ct values between groups. The Kappa test was initially applied to determine the agreement rate between the two PCR methods. The index was calculated using the Quickcalc GraphPad tool (https://www.graphpad.com/quickcalcs/kappa2). Statistical tests were applied with a 95% confidence interval.

## RESULTS

### Clinical-epidemiological characterization of the setting

From the 347 patients enrolled in the study, 332 (95.6%) had valid PCR results and were included in the analysis. From the 13 excluded patients, 7 had discordant (positive/negative) PCR results and eight had double invalid results (Figure 1). From the 332 patients that were included in the statistical analysis, 67 were positive for SARS-CoV-2 (based on PCR results) and the overall disease prevalence in the setting was 20.2%. As sample collection was performed during a four weeks span, the PCR positivity rate in the setting decreased continuously, from 35.5% (February 15th) to 2.8% (March 21th) (Figure 2A).

**Figure 2.**
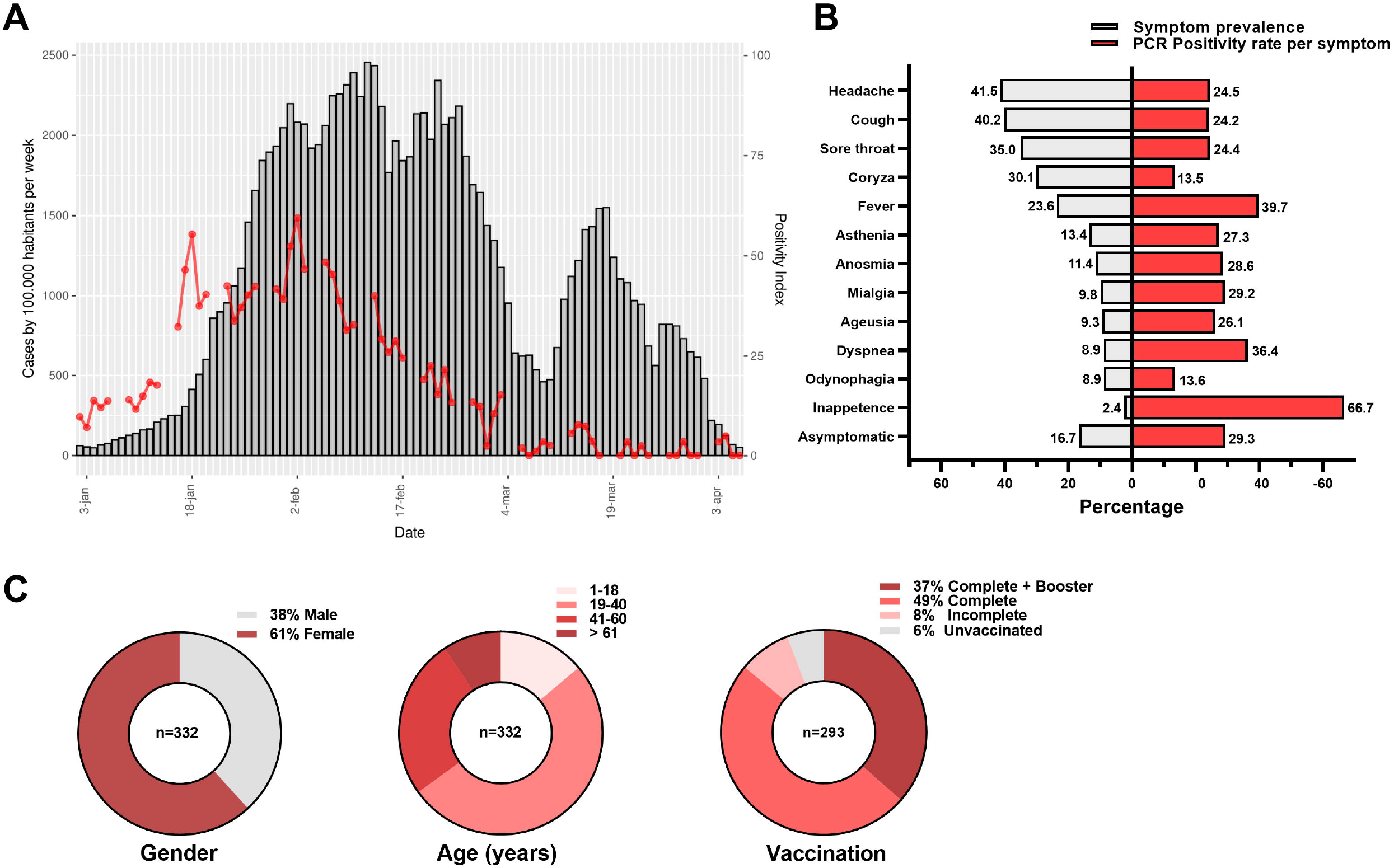
Epidemiological, clinical and demographic characterization of the cohort. **(A)** COVID-19 incidence in the city of Caruaru during the outbreak by the Omicron variant (left axis, gray bars) and positivity index of the tests performed at the city testing center (right axis, red line). **(B)** In gray, the prevalence of each symptom self-reported by the patients analyzed in the study and in red, the percentage of positive PCR tests for each reported symptom. **(C)** demographic features of the patients included in the study. The vaccination status was considered complete when the individual had two doses of Coronavac, AstraZeneca or Pfizer or one shot of Janssen and incomplete for cases with only one dose of Coronavac, AstraZeneca or Pfizer.

From the 332 individuals, 127 (38%) were male and 205 (62%) were female. The median age was 34 years-old (range: 1-96 years) and no significant differences were observed between the ages of the PCR-positive and negative and male/female individuals (p=0.526 and p=0.093, respectively). At the time of sample collection, 94% of the individuals enrolled in the study received at least one dose of a COVID-19 vaccine, 86% had a complete vaccination scheme (Coronavac/Sinovac, AstraZeneca, Pfizer or Janssen) (Figure 2B) and 84% of the individuals reported at least one COVID-19-related symptom (Figure 2C). The most common symptoms reported were headache, cough and coryza and the symptoms with the highest PCR positivity rate were inappetence (66.7%), fever (39.7%) and dyspnea (36.4%) (Figure 2C). Moreover, 16.7% of the patients reported to be asymptomatic, and from those, 29.3% tested positive by PCR.

### Validation of the PCR-combined reference method

Taking into consideration three possible outcomes (positive, inconclusive and negative) in the 347 collected samples, the Weighted Kappa index comparing the results from the two PCR methods was 0.885 (CI95% = 0.786-0.910), which is classified as “almost perfect agreement” (Supplementary Table 1). While analyzing the 67 positive samples, we observed a Pearson correlation rate of 0.872 between the Ct values of the E (Bio-manguinhos) and N1 (CDC) targets (Figure 3A).

**Figure 3.**
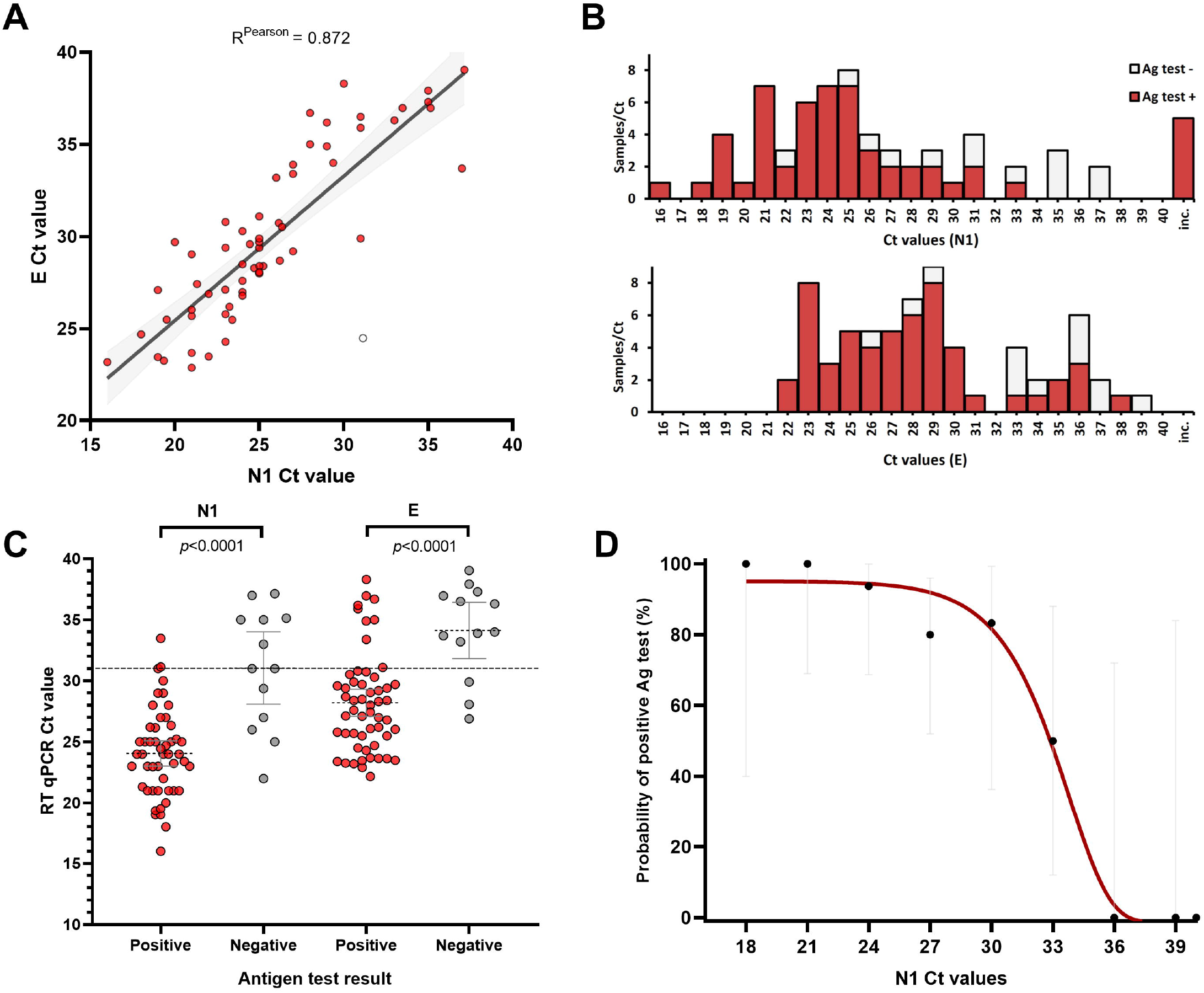
Comparison of the PCR and the rapid antigen test results. **(A)** correlation between the Ct values from the two PCR methods that were used combined as the reference method. The colorless datapoint represents the outlier sample **(B)** Distribution of Ct values and the proportion of positive samples in the rapid test. **(C)** Ct values grouped according to the rapid test result: Ag-positive samples had lower Ct values (higher viral load) than the Ag-negative samples. **(D)** Probability of positive Ag test according to the Ct values (N1 target) in groups of 3 (e.g. 16-18, 19-21). The curve was calculated by the non-linear asymmetric sigmoidal curve fit.

### Evaluation of the TR DPP® COVID-19 Ag test performance

We evaluated multiple performance parameters by comparing the results of the rapid antigen test and the PCRs. From the 67 patients that tested positive in the PCR, 54 also tested positive in the antigen test (overall sensitivity = 80.6%; CI95% = 69.11 -89.24%) and 2 out of the 265 PCR-negative patients were false-positive in the antigen test (specificity: 99.2%; CI95% = 97.3 – 99.1%). All antigen tests here performed presented a valid result. Moreover, we observed high positive and negative predictive values (96.4% and 95.3%, respectively), considering the prevalence of 20.2% in our setting. Further details can be found in Tables 1 and 2.

**Table 1.**

| **All samples** | **RT-qPCR** |  | **Total** |
| --- | --- | --- | --- |
|  | **Positive** | **Negative** |  |
| Ag Test + | 54 | 2 | 56 |
| Ag-Test - | 13 | 263 | 276 |
| Total | 67 | 265 | 332 |
| **Ct < 31 (E/Rp Bio)** |  |  |  |
| Ag Test + | 45 | - | - |
| Ag-Test - | 3 | - | - |
| Total | 48 | - | - |
| **Ct ≥ 31 (E/Rp Bio)** |  |  |  |
| Ag Test + | 9 | - | - |
| Ag-Test - | 10 | - | - |
| Total | 19 | - | - |
| **Ct < 31 (N1/N2 CDC)** |  |  |  |
| Ag Test + | 46 | - | - |
| Ag-Test - | 5 | - | - |
| Total | 51 | - | - |
| **Ct ≥ 31 (N1/N2 CDC)** |  |  |  |
| Ag Test + | 3 | - | - |
| Ag-Test - | 8 | - | - |
| Total | 11 | - | - |

**Table 2.**

|  | **Value** | **95% CI** |
| --- | --- | --- |
| **Overall sensitivity** | 80.60% | 69.11 - 89.24 |
| **E/RP** Ct < 31 | 93.75% | 82.80 - 98.69 |
| Ct ≥ 31 | 47.4% | 24.45 - 71.14 |
| **N1/CDC** Ct < 31 | 90.2% | 78.59 - 96.74 |
| Ct ≥ 31 | 27.3% | 6.02 - 60.97 |
| **Specificity** | 99.25% | 97.30 - 99.91 |
| **Disease prevalence** | 20.20% | - |
| **Positive Predictive Value** | 96.43% | 87.12 - 99.08 |
| **Negative Predictive Value** | 95.28% | 92.54 - 97.05 |
| **Accuracy** | 95.48% | 92.65 - 97.45 |

While these values are representative of the entire setting, we observed drastic differences in the test sensitivity across distinct PCR Ct values, with major drops in the Ct values higher than 31 (Figures 3B-D). Using a Ct value of 31 as cut-off, the test sensitivity dropped from 93.7% to 47.4% considering the N1 target and from 90.2% to 27.3% considering the E target (Table 2). Corroborating this finding, the average N1 Ct values for true positives and false negatives were 24.1 (CI95% = 23-25.1) and 31 (CI95% = 28.1-34), respectively, and for the E target were 28.2 (CI95% = 27.1-29.3) and 34.1 (CI95% = 31.2-36.4), respectively (Figure 3C).

### Genomic characterization of SARS-CoV-2 and phylogenomic data analysis

We were able to sequence the SARS-CoV-2 genome of 40 out of 67 positive cases, with an average coverage and depth of 96.4% (range: 87.7-99.1%) and 245.1 (range: 72.7-948.5), respectively. From the 40 samples sequenced, 32 (80%) had a coverage higher than 95%. Additionally, other 17 samples with higher Ct values and ineligible for sequencing, were characterized with the multiplex qPCR variant-inference protocol. All the 57 successfully characterized samples (85% from all positives) were assigned as Omicron, revealing an Omicron-dominant setting, which is consonant with the epidemiological scenario from the region. The details of each positive case can be found in Supplementary Table 2.

In the phylogenetic analysis, the Nexstrain sampling returned 902 sequences from Pernambuco state, 222 from Brazil, 84 from South America, 50 from the globe and 42 from the output group. The phylogenetic tree encompasses 1340 genomes, 1300 from nexstrain sampling and 40 from the samples included in the study’s cohort. The phylogenetic analysis supports the lineage signature of pangolin tool (Figure 4), in which the sequenced genomes are allocated on BA.1 and BA.1-like lineage clades, while showing a wide genomic diversity of the sequenced samples within those clades.

**Figure 4:**
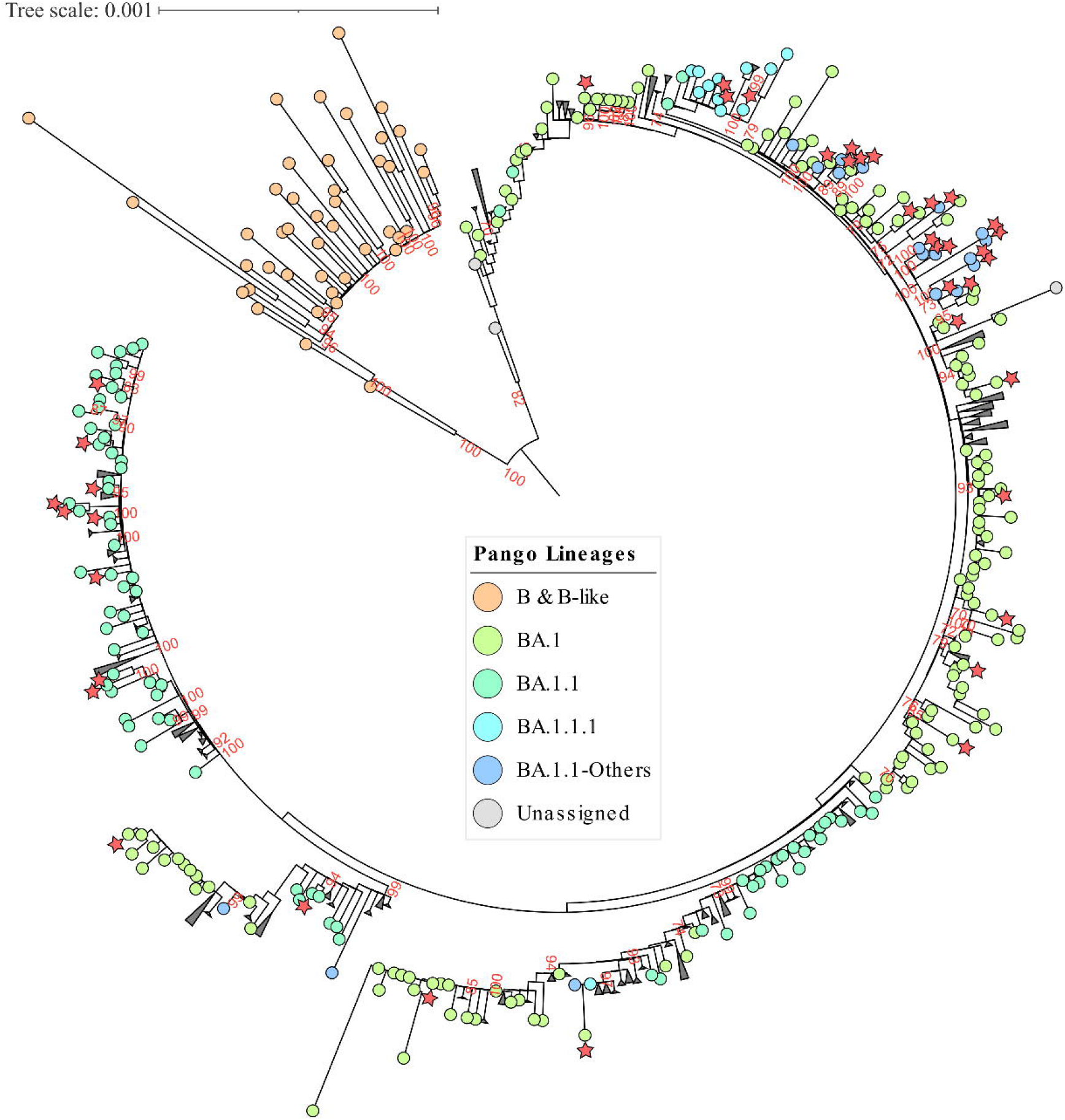
Maximum Likelihood phylogenetic tree focused on Omicron lineage. Branch support represented by ultrafast bootstrap. The genomes from the present study are marked with red stars.

## DISCUSSION

The current study investigated the performance of the DPP® SARS-CoV-2 Antigen test, which is widely used in the testing program from the public healthcare system in Brazil. We evaluated this test in a primary healthcare setting, including both symptomatic and asymptomatic individuals that reached the service for COVID-19 testing during the 2022 Omicron wave in the country.

The overall test sensitivity and specificity (80.6 and 99.2%, respectively) were within the recommended by the World Health Organization (WHO, 2021) and other organizations, such as the European Commission (ECDC, 2022). As previously reported for other antigen tests (Michelena *et al*., 2022; Albert *et al*., 2021; Alemany *et al*., 2021; Korenkov *et al*., 2021), the sensitivity reduced dramatically in samples with lower viral loads. Of importance, the overall sensitivity observed in the current study was lower than the reported in the product specifications (sensitivity: 90.3 and specificity: 98.8%, www.bio.fiocruz.br). However, this divergence in test sensitivity is probably due to the differences in the study design, which results in distinct distribution of Ct values (viral load) between the two settings. The results reported by the manufacturer included only symptomatic individuals with up to nine days from symptoms onset, and 79% of the positive samples had a Ct value lower than 25 (high viral load), while in our study only 48% of the positive cases were within this group. Nevertheless, when analyzing only samples within the Ct < 25 group, we observed a test sensitivity of 96.6%, which is similar to the reported by the manufacturer’s (98%) for this particular subset. By prospectively analyzing unselected patients in a real-life study (all-comers), we were able to avoid bias towards the selection of samples that are more likely to carry a higher viral load and thus, allowing a more realistic evaluation of the test.

The combined use of two distinct diagnosis-approved PCR protocols as reference methods increased the precision of the data obtained, avoiding bias caused by eventual errors during the execution of the PCR technique. Furthermore, the study brings two important features that must not be overlooked: i) all samples were evaluated fresh at point-of-care for the rapid antigen test, which is more representative of the routine diagnosis conditions than retrospective frozen samples, and ii) differently from studies carried out in late 2020/yearly 2021, it evaluates the test in a cohort with high vaccination coverage, which might influence the viral load and the duration of the active infection. On the other hand, one important limitation of the study is that, due to the fast decrease in the number of cases, only 67 positive patients were assessed, reducing the statistical resolution of the analysis in positive subgroups.

At a disease prevalence of 20.2%, the rapid antigen test performed with both positive and negative predictive values higher than 95%, which supports its use for COVID-19 diagnosis in high incidence settings without the need of confirmatory PCR results. However, it is important to stress that: i) these data should not be extrapolated to other contexts (e.g. testing in hospital admissions) and ii) predictive values are largely influenced by the disease prevalence and the dynamics of the epidemiologic scenario must be taken into consideration for interpreting the tests (WHO, 2021).

The Omicron carry numerous mutations and have marked clinical and biological differences when compared to the early 2020 strains and other VOCs (Viana *et al*., 2022; Jassat *et al*., 2022). Indeed, due to its substantial capability of evading neutralizing antibodies acquired from vaccines or previous infections, Omicron has been suggested to be classified as a distinct serotype from the other strains (Simon-Loriere *et al*., 2022). Similarly to most antigen rapid tests, the DPP® SARS-CoV-2 Antigen detects the nucleocapsid protein and has been developed and validated before the surge of the Omicron variant of concern. Taking into consideration that Omicron has at least four synapomorphic mutations in the N gene (P13L, del31/33, R203K and G204R)(Wang *et al*., 2022), it is of utmost importance to re-evaluate the performance of antigen tests in clinical samples from patients infected with this variant. Here, we were able to confirm the Omicron (BA.1/BA.1.1) lineage in all samples eligible for whole-genome sequencing or multiplex PCR (85% of all positives).

Recent studies reported that the Omicron variant did not impact the limit of detection (LOD) of antigen tests in experiments with cultured virus (Stanley *et al*., 2022; Raich-Regué *et al*., 2022) nor its performance at clinical level (Galliez *et al*., 2022; Michelena *et al*., 2022). While these studies evaluated other models of antigen tests, their findings are in line with our data regarding the clinical evaluation of the DPP® SARS-CoV-2 test. Of interest, since the BA.2 subvariant carries the exact same mutations in the nucleocapsid as the BA.1, similar antigen test patterns would be expected for this lineage. However, further studies will be required to evaluate the performance of antigen tests with the newly identified BA.4 and BA.5 sub lineages, that carry a distinct S413R mutation in the N gene (www.outbreak.info).

Altogether, the data obtained here supports the maintenance of the rapid antigen test sensitivity for the BA.1 and BA.1.1 Omicron lineage and underscores the reliability of the test for COVID-19 diagnosis in high transmission settings.

## Data Availability

All data produced in the present study are available upon reasonable request to the authors

## ACKNOWLEDGMENTS

We are thankful to the healthcare professionals from the Centro Municipal de Testagem de Caruaru for working along with our team to make this work possible. We also appreciate the support of the Fiocruz COVID-19 Genomic Surveillance Network (http://www.genomahcov.fiocruz.br/).

