## Supplementary material for "Real-life evaluation of a rapid antigen test (DPP® SARS-CoV-2 Antigen) for COVID-19 diagnosis of primary healthcare patients, in the context of the Omicron-dominant wave in Brazil": Supp Tables

| **Supplementary Table 1.** Weighted Kappa test PCR N1/N2(CDC) and Duplex SARS-CoV-2 E/RP Molecular Kit. | | | | | | |  |
| --- | --- | --- | --- | --- | --- | --- | --- |
|  |  | **N1/N2** | | |  | **Legend:** | **Valid** |
|  |  | **Positive** | **Inconclusive** | **Negative** | **Total** |  | **Invalid** |
| **E/RP** | **Positive** | 62 | 5 | 5 | 72 |  |  |
|  | **Inconclusive** | 0 | 8 | 0 | 8 |  |  |
|  | **Negative** | 2 | 8 | 257 | 267 | **Weighted Kappa:  0.885** |  |
|  | **Total** | 64 | 21 | 262 | 347 |  |  |

**Supplementary Table 2.** Description of SARS-CoV-2 Positive samples submitted to whole-genome sequencing.

| Gender | Vaccination | Antigen test | Ct (N1) | Ct (E) | Depth | Coverage | GISAID ID | Lineage_org | Lineage_up |
| --- | --- | --- | --- | --- | --- | --- | --- | --- | --- |
| Female | complete | Positive | 21 | 22,9 | 699.05 | 98,69 | EPI_ISL_11894686 | BA.1 | Omicron (BA.1-like) |
| Male | complete | Positive | 21 | 25,7 | 577.2 | 96,43 | EPI_ISL_11894687 | BA.1 | Omicron (BA.1-like) |
| Female | complete | Positive | 23 | 25,8 | 809.25 | 97,76 | EPI_ISL_11894688 | BA.1 | Omicron (BA.1-like) |
| Male | complete | Positive | 24 | 27 | 151.77 | 98,28 | EPI_ISL_11894703 | BA.1 | Omicron (BA.1-like) |
| Male | complete | Positive | 19 | 23,47 | 948.45 | 99,13 | EPI_ISL_11894689 | BA.1 | Omicron (BA.1-like) |
| Male | complete | Positive | 25 | 29,4 | 126.52 | 92,7 | EPI_ISL_13222862 | BA.1 | Omicron (BA.1-like) |
| Female | booster | Positive | 25 | 28 | 129.77 | 91,24 | EPI_ISL_13222863 | BA.1.1 | Omicron (BA.1-like) |
| Female | complete | Positive | 27 | 29,2 | 169.59 | 93,27 | EPI_ISL_13222864 | BA.1 | Omicron (BA.1-like) |
| Female | complete | Positive | 22 | 23,5 | 331.0 | 96,59 | EPI_ISL_11894690 | BA.1 | Omicron (BA.1-like) |
| Female | complete | Positive | 16 | 23,2 | 465.15 | 95,53 | EPI_ISL_11894691 | BA.1 | Omicron (BA.1-like) |
| Male | unvaccinated | Positive | 27 | 33,4 | 127.35 | 87,87 | EPI_ISL_13222865 | BA.1 | Omicron (BA.1-like) |
| Female | incomplete | Positive | 19 | 27,1 | 775.74 | 98,59 | EPI_ISL_11894692 | BA.1 | Omicron (BA.1-like) |
| Male | complete | Positive | 24 | 30,3 | 195.1 | 97,37 | EPI_ISL_11894693 | BA.1 | Omicron (BA.1-like) |
| Male | booster | Positive | 24 | 27,6 | 172.3 | 98,01 | EPI_ISL_11894701 | BA.1.1 | Omicron (BA.1-like) |
| Female | complete | Positive | 23 | 30,8 | 168.79 | 98,33 | EPI_ISL_11894694 | BA.1 | Omicron (BA.1-like) |
| Female | complete | Positive | 28 | 36,7 | 72.73 | 87,8 | EPI_ISL_13222866 | BA.1.1 | Omicron (BA.1-like) |
| Female | booster | Positive | 21 | 26,03 | 200.91 | 98,33 | EPI_ISL_11894702 | None | Omicron (Unassigned) |
| Male | booster | Positive | 23 | 29,4 | 185.46 | 98,33 | EPI_ISL_11894695 | BA.1.1 | Omicron (BA.1-like) |
| Male | booster | Positive | 25 | 29,7 | 156.44 | 96,98 | EPI_ISL_11894696 | BA.1 | Omicron (BA.1-like) |
| Male | complete | Positive | 18 | 24,7 | 171.92 | 98,28 | EPI_ISL_11894697 | BA.1.1 | Omicron (BA.1-like) |
| Female | booster | Positive | 20 | 29,7 | 168.9 | 98,15 | EPI_ISL_11894698 | BA.1 | Omicron (BA.1-like) |
| Female | booster | Negative | 27 | 33,9 | 79.28 | 87,95 | EPI_ISL_13222867 | BA.1 | Omicron (BA.1-like) |
| Male | complete | Positive | 24 | 28,5 | 152.36 | 98,14 | EPI_ISL_11894699 | BA.1.1 | Omicron (BA.1-like) |
| Female | booster | Negative | 22 | 26,9 | 192.66 | 98,29 | EPI_ISL_11894700 | BA.1.1 | Omicron (BA.1-like) |
| Female | booster | Positive | 23 | 24,3 | 200.37 | 98,27 | EPI_ISL_11894704 | BA.1 | Omicron (BA.1-like) |
| Female | booster | Positive | 24 | 26,8 | 201.28 | 98,18 | EPI_ISL_11894705 | None | Omicron (Unassigned) |
| Female | complete | Negative | 26 | 33,2 | 83.07 | 87,74 | EPI_ISL_13222868 | BA.1 | Omicron (BA.1-like) |
| Female | complete | Positive | 21 | 23,7 | 219.75 | 98,33 | EPI_ISL_11894706 | BA.1 | Omicron (BA.1-like) |
| Female | complete | Positive | 19,35 | 23,27 | 188.64 | 98,34 | EPI_ISL_11894713 | BA.1 | Omicron (BA.1-like) |
| Female | complete | Positive | 25,23 | 28,4 | 124.51 | 97,57 | EPI_ISL_11894716 | BA.1 | Omicron (BA.1-like) |
| Male | unvaccinated | Positive | 26,16 | 30,74 | 187.95 | 97,51 | EPI_ISL_11894711 | BA.1.1 | Omicron (BA.1-like) |
| Male | booster | Positive | 24,7 | 28,3 | 155.72 | 96,83 | EPI_ISL_11894715 | BA.1 | Omicron (BA.1-like) |
| Female | booster | Positive | 25,01 | 28,4 | 175.12 | 97,43 | EPI_ISL_11894717 | BA.1.1 | Omicron (BA.1-like) |
| Female | complete | Positive | 23,4 | 25,49 | 128.35 | 98,09 | EPI_ISL_11894712 | BA.1.1 | Omicron (BA.1-like) |
| Female | complete | Negative | 25 | 28,07 | 174.42 | 98,33 | EPI_ISL_11894707 | BA.1.1 | Omicron (BA.1-like) |
| Female | complete | Positive | 23,23 | 26,2 | 177.27 | 98,29 | EPI_ISL_11894708 | BA.1.1 | Omicron (BA.1-like) |
| Female | booster | Positive | 26,35 | 30,53 | 88.55 | 94,91 | EPI_ISL_13222869 | BA.1 | Omicron (BA.1-like) |
| Female | booster | Positive | 21,32 | 27,43 | 133.38 | 98,36 | EPI_ISL_11894714 | BA.1.1 | Omicron (BA.1-like) |
| Male | booster | Positive | 19,52 | 25,5 | 160.19 | 98,76 | EPI_ISL_11894710 | BA.1 | Omicron (BA.1-like) |
| Male | complete | Positive | 24,44 | 29,59 | 176.25 | 97,92 | EPI_ISL_11894709 | BA.1.1 | Omicron (BA.1-like) |
